# Lumbar puncture and the diagnosis of aneurysmal subarachnoid haemorrhage – a ten-year service evaluation

**DOI:** 10.64898/2026.09.02.26362072

**Authors:** Jamie Talbot, Daniel Lashley

## Abstract

**Objectives:** Aneurysmal subarachnoid haemorrhage is a life-threatening emergency that commonly presents with a sudden, severe headache. The diagnosis is usually confirmed on CT imaging, however a small subset of patients demonstrate normal imaging, particularly if presentation is delayed – in which case spectrophotometric analysis of cerebrospinal fluid (CSF) is undertaken. Nevertheless, thunderclap headache is a common presentation and the vast majority of investigated patients do not have aneurysmal subarachnoid haemorrhage. This ten-year service evaluation at a tertiary neurosurgery centre aims to estimate the utility of lumbar puncture in the diagnostic work-up of these patients.

**Methods:** All instances of CSF spectrophotometry between 01/01/2011 and 31/12/2020 were evaluated. Clinical records of patients with positive spectrophotometry were analysed. A second method identified all coded diagnoses of non-traumatic subarachnoid haemorrhage over the same time period, with review of clinical records in cases of negative imaging.

**Results:** Of 2158 instances of CSF spectrophotometry performed at the tertiary neurosurgery centre, four aneurysms were identified and treated by coiling or clipping, resulting in a number needed to treat of 540 patients to diagnose one case of aneurysmal subarachnoid haemorrhage. Of 648 patients with a coded diagnosis of non-traumatic subarachnoid haemorrhage (including local patients and transfers from satellite hospitals), 27 patients displayed initial negative imaging, of which 9 were found to have aneurysms and underwent neuro-intervention. Lumbar puncture was instrumental in the diagnosis and led to intervention in three patients identified by this method.

**Conclusion:** CSF analysis resulted in diagnosis and intervention for missed (CT-negative) aneurysmal subarachnoid haemorrhage in approximately 1/540 tested patients. The estimated yield in a real-world setting may be useful for clinicians or provide a basis for discussion when counselling patients on benefit and risk.

## 1. Background

Acute subarachnoid haemorrhage (SAH) due to rupture of a saccular aneurysm is a life-threatening illness associated with high mortality and morbidity. In patients presenting with suspected SAH, computed tomography (CT) of the brain is the primary investigation of choice, with a high sensitivity for detecting subarachnoid haemorrhage immediately after symptom onset (Dubosh et al., 2016; Edlow et al., 2009; Perry et al., 2011; American College of Emergency Physicians Clinical Policies Subcommittee (Writing Committee) on Acute Headache: et al., 2019). However, over time the sensitivity declines, with estimated sensitivity of only 50% after 1 week (Morgenstern et al., 1998). Spectrophotometric analysis of the cerebrospinal fluid (CSF) is commonly performed in the event of a negative CT scan, with CSF bilirubin detectable for up to 2 weeks after the onset of headache (Cruickshank et al., 2008).

Lumbar puncture is invasive and time-consuming, and its utility in the diagnosis of subarachnoid haemorrhage has been subject to significant scrutiny over the years (Boesiger and Shiber, 2005; Brunell et al., 2013; Foot and Staib, 2001; O’Neill et al., 2005; Perry et al., 2008). Detection rates of SAH have notably improved with the development of third generation CT scanners with multiple rows of detectors, with one landmark Canadian study reporting 100% sensitivity and specificity when performed within 6 hours of headache onset (Perry et al., 2011). Some recent studies have suggested negligible benefit of lumbar puncture in identifying missed cases of aneurysmal SAH, with two retrospective studies with a pooled total of 587 patients identifying no instances where lumbar puncture led to neuro-intervention of an aneurysm in patients with negative scans (Brunell et al., 2013; Gee et al., 2012).

There is considerable variation in clinical practice of physicians managing suspected subarachnoid haemorrhage with limited clinical guidance. In the UK, the National Institute of Clinical Excellence (NICE) draft guide advises consideration of lumbar puncture in all patients with suspected SAH who undergo CT head imaging over six hours after the onset of symptoms (https://www.nice.org.uk/guidedance/GIDNG10097/documents/draft-guideline). The guidelines advise that most patients with SAH experience a sudden, severe ‘thunderclap’ headache and may have associated symptoms such as neck pain or stiffness, photophobia, vomiting or altered neurology, but does not otherwise present a framework for ratio-nalising the use of lumbar puncture. In certain cases of severe or thunderclap headache, especially those with more benign-sounding features or with relative contra-indications or challenges to lumbar puncture, patients are sometimes counselled about the anticipated diagnostic benefit of further diagnostic work-up and decisions are made jointly about whether to proceed. Nevertheless, these risk-benefit estimates are prone to significant error and variation, based on clinical studies spanning several decades with numerous discrepancies in imaging technology, inclusion criteria and the definition of ‘thunderclap’ headache.

This 10-year retrospective service evaluation examines realworld outcomes in patients undergoing diagnostic work-up for subarachnoid haemorrhage at a tertiary neurosurgery centre in the UK - serving a population of around 1.2 million people for neurosurgical care, and approximately 450,000 for neurology/general medical care. It seeks to evaluate the utility of lumbar puncture in the diagnosis of aneurysmal SAH in the setting of routine clinical practice and hopes to provide a modern-day estimate for the diagnostic yield of this approach, as well as examining clinical features of patients with confirmed aneurysmal subarachnoid haemorrhage with initial negative CT scans. We also provide some observational data providing insights into the prevalence of ‘sentinel’ headaches - patients presenting with acute, severe headaches in the weeks prior to aneurysmal SAH.

## 2. Methods

This retrospective study was conducted at Derriford Hospital, Plymouth, a tertiary neurosurgery centre in the South West of England, UK. It was registered and approved locally as a service evaluation, with relevant discussion with the research and development department to confirm the study did not meet the requirements for ethics approval. Data was collected between 2021 and 2022.

Data was acquired by two methods: Firstly, all patients who underwent CSF spectrophotometry at Derriford Hospital between 01/01/2011 and 31/12/2020 were identified. In patients in whom bilirubin was detected in the CSF, paper and electronic hospital records were reviewed and data relating to clinical features, diagnosis, investigations and outcomes was collected.

The second method identified all patients with a coded primary diagnosis of non-traumatic subarachnoid haemorrhage over the same time period. In contrast to the first method, which identified patients who directly presented to the tertiary neurosurgery centre, this also included patients transferred from satellite hospitals, including on the basis of CT-negative, xanthochromia-positive results. Imaging reports were examined and classified on the basis of the presence or absence of intracranial haemorrhage on initial CT. In scans that were secondarily reported by senior radiologists, the secondary report was used for classification. Scans with ambiguous reporting (e.g. ‘possible’, ‘may represent’) were classified as negative, whilst those with positive reporting (e.g. ‘definite’, ‘probable’) were classified as positive. In patients without evidence of SAH on cranial imaging, hospital records were scrutinised for further information.

## 3. Results

### 3.1 Spectrophotometric analysis of CSF

Between 01/01/2011 and 31/12/2020, CSF bilirubin sampling was undertaken on 2158 occasions in 2075 different patients. Of these, CSF bilirubin was detected on 45 occasions (2.1%), in the same number of patients. An alternative/nonspontaneous cause was identified in 19/45 patients, including high CSF protein (1/19), meningoencephalitis (8/19), hepatic encephalopathy (1/19), hypertension (1/19), cortical vein thrombosis (1/19), reversible cerebral vasoconstriction syndrome (1/19), prior traumatic lumbar puncture (3/19), trauma (1/19), cocaine use (1/19), and viral illness (1/19). Twenty-three patients underwent digital subtraction angiography, of which 2 patients were found to have aneurysms and underwent intervention (1 clipping; 1 coiling). Two additional patients underwent intervention (1 clipping; 1 coiling) on the basis of CT angiography alone, one of whom was already under surveillance for a known MCA aneurysm.

### 3.2 Primary coded diagnosis of non-traumatic subarachnoid haemorrhage

There were 648 coded diagnoses of non-traumatic subarachnoid haemorrhage between 01/01/2011 and 31/12/2020. Twenty seven of the 648 patients (4.2%) had no evidence of intracranial haemorrhage on initial CT or other brain imaging. Of these, 13/27 patients had CSF sampled at the regional neurosurgery centre, with CSF bilirubin detected in 12/13 patients – twelve of these patients underwent digital subtraction angiography, with 2 undergoing intervention (1 coiling; 1 clipping; the same patients as identified above). Of the remaining 14 patients, 8 were referred from satellite hospitals, of which 7/8 were transferred on the basis of negative CT/positive spectrophotometry. Subarachnoid haemorrhage was diagnosed in the remaining patients on the basis of clinical features (e.g. hydrocephalus, vasospasm), usually with identification of an aneurysm or due to subsequent bleeding. A total of 9/27 patients with negative imaging underwent intervention for an aneurysm during the study period (7 coiling, 2 clipping). Lumbar puncture was instrumental in the diagnosis of aneurysmal subarachnoid haemorrhage and led to intervention in a total of 3 patients. Figure 1 depicts a flowchart illustration of these results.

**Figure 1.**
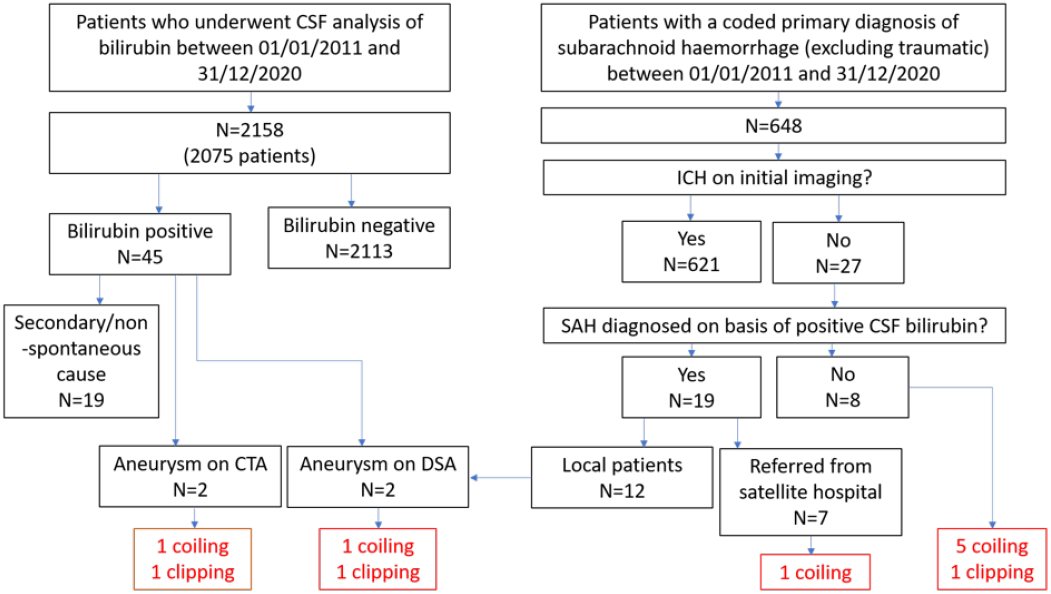
Flowsheet of audit findings. CSF spectrophotometry performed at the tertiary centre resulted in four instances of neuro-intervention out of 2158 instances of sampling. Of 648 patients with a coded diagnosis of non-traumatic SAH, lumbar puncture was instrumental in the diagnosis of SAH in 19 patients, with three going on to receive neuro-intervention. In total, lumbar puncture was instrumental in the diagnosis of aneurysmal SAH in five patients across the region who went on to receive neuro-intervention. CTA=CT angiogram, DSA=digital subtraction angiography, ICH=intracranial haemorrhage.

### 3.3 Clinical features of patients demonstrating CT-negative subarachnoid haemorrhage

The clinical features of 11 patients with initial negative CT who received intervention for an aneurysm was analysed. Seven (64%) were female. The mean age was 57 (range 34-86). Five of 11 patients presented with a typical thunderclap headache. Two patients presented with headaches in conjunction with III nerve palsy, one of whom deteriorated in hospital with severe headaches and vomiting and repeat CT showed SAH. Two patients presented with gradual onset headaches, both with meningism, vomiting and high pressure features. One patient presented with neck pain, who deteriorated with confusion, meningism, vomiting and hypertension. Another presented with a seizure with severe headache and meningism. Of the five patients who presented with thunderclap headache, the mean time until presentation was 3.4 days (range 1-7). Three of the five patients displayed vomiting, 4/5 had neck stiffness, 3/5 had meningism, 1/5 had confusion.

### 3.4 Imaging for recent ‘sentinel’ headaches

Sentinel, or warning headaches in the days or weeks prior to subarachnoid haemorrhage, are said to occur in as many as 15-60% of patients with spontaneous SAH (Polmear, 2003; Verweij et al., 1988). A crude method of assessing the occurrence of a warning headache in the prior 6 months was applied. In the literature, these warning headaches are described as severe (Polmear, 2003), and therefore it is reasonable to expect that patients will present to hospital and receive cranial imaging. Patients with a coded primary diagnosis of non-traumatic SAH were analysed for undergoing CT or other cranial imaging in the 6 months prior to SAH. Of 648 patients, 273 resided outside the catchment area of the regional neurosurgery centre and were excluded, leaving 374 local patients presenting with SAH. Thirty-nine of 374 patients (10.4%) had received cranial imaging within 6 months of their presentation, of which 4/374 (1.1%) were performed for acute headaches. Of these, all were negative for acute bleeding. Two of the 4 patients underwent investigation with lumbar puncture at the time, negative for bilirubin in both cases. Of the remaining patients with recent imaging, 9/273 (3.3%) had recently suffered a SAH, 7/273 (2.6%) were receiving aneurysm surveillance, and 2/273 (0.7%) had been recently diagnosed with an aneurysm, one of whom was awaiting intervention.

## 4. Discussion

This study spanning 10 years reveals that in patients presenting to the regional neurosurgical centre or transferred from a satellite hospital with suspected SAH, lumbar puncture was instrumental in the diagnosis of aneurysmal SAH in only five patients who went on to receive neuro-intervention. Analysis of CSF within the tertiary referral centre was performed on 2158 occasions over 10 years, of whom only four patients (0.19%) with positive spectrophotometry were found to have an aneurysm and received intervention, producing a number needed to treat (NNTT) of approximately 540 to diagnose and treat one aneurysmal SAH.

It is not within the scope of this article to address the costbenefit analysis of lumbar puncture in the diagnostic work-up of aneurysmal subarachnoid haemorrhage. Neurological injury resulting from this diagnosis is responsible for significant morbidity as well as extensive societal costs relating to healthcare usage, lost productivity and requirement for social care input, and therefore the high number of patients screened to detect a single case may be financially and morally justified. Instead, this real-world service evaluation is useful in estimating the yield of lumbar puncture in the contemporary work-up for aneurysmal SAH, which relates the individual practice of multiple different specialists working together within the remit of the tertiary neurosurgical centre. Whilst the outcomes will be influenced by a number of factors at both a clinical and service level, the results may well be generalisable to secondary care centres both in the UK and abroad, with or without local neurosurgical provision.

When considering the diagnosis of aneurysmal subarachnoid haemorrhage in patients that present with acute, severe headaches with negative imaging, the decision to proceed to lumbar puncture may be driven by a range of factors ranging from clinical symptoms and signs, timing of imaging, patient preference, availability of beds and skilled practitioners, contraindications or challenges relating to the procedure, concurrent medications such as warfarin or clopidogrel which need to be stopped a number of days prior to procedure with its own inherent risks, as well as individual thresholds for tolerating risk. Physicians are routinely expected to counsel patients on risk in clinical practice – by providing accurate information, patients are able to adequately weigh up clinical decisions to make joint decisions about their care. Despite numerous clinical studies on the subject of aneurysmal subarachnoid haemorrhage published over a number of decades, risk-benefit estimates are still often hard to extrapolate in real-world practice which continues to hamper these discussions.

NICE’s draft guidelines advise that most patients with SAH experience a sudden, severe ‘thunderclap’ headache and may have associated clinical features, but does not otherwise present a framework for rationalising the use of lumbar puncture and tacitly advocates use of one’s own judgement in these situations. It is conventional (and stipulated in NICE’s draft guidance) to consider lumbar puncture in all patients with suspected SAH who demonstrate negative imaging if they receive their CT scan greater than six hours after the onset of headache, and this service evaluation demonstrated that all such patients who were eventually diagnosed with aneurysmal SAH underwent imaging at least one day after the onset of headache, with the caveat of significant selection bias. In patients with aneurysmal SAH and negative CT imaging, thunderclap headache was re-ported in 5/11 patients, in which the majority had neck stiffness (80%), vomiting (60%) and meningism (60%). The absence of these clinical signs would therefore be reassuring in a patient with suspected SAH and negative CT imaging.

Our analysis incorporated a crude method to assess for the occurrence of recent sentinel headache in the 6 months prior to their diagnosis of non-traumatic subarachnoid haemorrhage. Surprisingly, only a small fraction of patients with this coded diagnosis within the hospital’s catchment (1.1%) had received recent imaging for a headache. Two patients had undergone work-up with lumbar puncture and CSF spectrophotometry which failed to identify xanthochromia in either case. Although the methodology is unsophisticated, these findings suggest that sentinel headaches are either less common than typically suggested or infrequently result in patients attending hospital or undergoing cranial imaging. Furthermore, a previous negative xanthochromia result should not necessarily be a reassurance when investigating for suspected aneurysmal SAH.

As a service evaluation, the outcomes are a reflection of local clinical practice which will unavoidably display regional and international discrepancies. It will also be biased by its non-inclusion of patients that failed to undergo lumbar puncture, for example due to technical difficulty or patient choice. Nevertheless, the findings are a summation of clinical practice within a tertiary hospital over a relatively long period of time, and therefore provide a reasonable real-world estimate of the utility of lumbar puncture in the diagnosis of aneurysmal SAH. The dual method of patient selection, both by instances of CSF spectrophotometry and coded diagnosis of SAH, would expect to identify all cases of aneurysmal subarachnoid haemorrhage as well as secondary causes and perimesencephalic SAH, however some cases could have been missed. The latter selection method (via coded diagnosis) in particular failed to identify two patients with aneurysmal SAH identified by CSF spectrophotometry, and was additionally confounded by significant heterogeneity, containing cases of cerebral amyloid angiopathy and subdural haemorrhage with subarachnoid extension. The classification of subarachnoid haemorrhage on the basis of consultant radiologist reporting is also problematic in the context of subtle or equivocal scan findings – as a result, this study may underestimate the incidence of CT-negative SAH and be less generalisable to settings such as district hospitals.

## 5. Conclusion

Lumbar puncture is conventionally performed in patients presenting with suspected subarachnoid haemorrhage without radiological evidence of haemorrhage on CT imaging. This analysis found that the procedure resulted in intervention in only a small fraction of patients, with an estimated NNT of 540 to diagnose one aneurysmal SAH. Awareness of the diagnostic utility of lumbar puncture in real-world practice may help guide clinical decisions about use of this test and provide a basis for patient discussions when counselling on benefit and risk.

## Data Availability

All data produced in the present work are contained in the manuscript

